# THE EFFECT OF BARICITINIB USAGE ON THE CLINICAL AND BIOCHEMICAL PROFILES OF COVID-19 PATIENTS- A RETROSPECTIVE OBSERVATIONAL STUDY

**DOI:** 10.1101/2021.08.11.21261760

**Authors:** Anoop Amarnath, Ananya Das, Venakata Sai Shashank Mutya, Irfan Ibrahim

## Abstract

**Introduction:** Coronaviruses typically cause influenza like illness which progresses to ARDS along with other systemic manifestations. India is experiencing its second wave with a huge surge in the number of cases exponentially causing huge impact on health care infrastructure and the demand supply chain. As a result several new modalities have been used, one of which is the use of remdesivir and baricitinib . Hence this study is aimed at finding out the clinical and biochemical profile of the patients who have received the combination

**Materials and Methods:** All the patients who have received the combination meeting the inclusion and exclusion criteria have been included in the study. A total of 31 participants were included and their records were retrospectively analyzed.

**Results:** There was a significant reduction in the oxygen requirement, CRP and IL-6 levels with p values<0.05. However, in the non-survivors group, there was no statistically significant reduction. Serial monitoring of NLR ratio showed increase towards the fifth day, especially in the non-survivor group it was as high as 41.24. The mortality rate was found to be 10% and the cause being secondary sepsis in all of them.

**Conclusion:** The ACTT-2 trail has proved the efficacy of the use of the remdesivir and baricitinib combination with mortality benefit. In our study we found similar results which was well co-related with clinical and biochemical parameters like CRP and IL-6 especially in people with co-morbidites.

## INTRODUCTION

Coronaviruses are enveloped non segmented positive sense RNA viruses belonging to the family Coronaviridae and broadly distributed in humans and other mammals. Although most human corona virus infection are mild, the epidemics of the two beta coronaviruses, Severe Acute Respiratory Syndrome- Coronavirus(SARS-CoV) IN 2003 AND Middle East Respiratory Syndrome( MERS-CoV) In 2012 have caused more than 10,000 cumulative cases in the past two decades with mortality rates of 10% for SARS-CoV and 37% for MERS-CoV. Severe Acute Respiratory Syndrome – coronavirus 2 SARS-CoV emerged as a zoonotic in 2019 and is the causative agent for COVID-19^[1]^. Exposure to SARS-CoV can result in a range of clinical outcomes varying from asymptomatic infection to ARDS and death. SARS-CoV has spread globally and on March 11, 2020 it was declared as a pandemic by WHO ^[1]^.

The COVID-19 infection typically causes influenza like illness in the early stage with gradual progression to ARDS. But of late new symptoms have been identified namely: myalgia, diarrhea, conjunctivitis, rashes on the body, sore throat and headache. The COVID-19 virus has undergone various mutations which has led to vaccine inefficacy, increasing transmissibility and mortality. The notable variants are UK variant( ALPHA ), Brazil variant (GAMMA ), South African variant ( BETA) and Indian variant ( DELTA )^[2]^

Currently, India is experiencing its second wave of COVID-19 due to these mutations with a surge in the number of cases exponentially causing a huge impact on health care infrastructure and the demand and supply chain. The state of Karnataka especially Bangalore was the worst hit in the second wave with more than 5 lakh reported cases. The increase in the number of cases threw a challenge to the treating clinicians to explore newer modalities of treatment in view of replenishing stocks of medicines and to reduce mortality. Several newer modalities were published and practiced in the Western world when they had surge in cases. One among the newer modalities is the use Baricitinib and Remdesivr combination.

Remdesivir is an intravenous nucleotide prodrug of an adenosine analog. Remdesivir binds to the viral RNA-dependent RNA polymerase and inhibits viral replication through premature termination of RNA transcription. It has demonstrated in vitro activity against SARS-CoV-2^[3]^. Remdesivir can cause gastrointestinal symptoms (e.g., nausea), elevated transaminase levels, an increase in prothrombin time (without a change in the international normalized ratio), and hypersensitivity reactions. Liver function tests and prothrombin time should be obtained in all patients before remdesivir is administered and during treatment as clinically indicated. Remdesivir may need to be discontinued if alanine transaminase (ALT) levels increase to >10 times the upper limit of normal and should be discontinued if an increase in ALT level and signs or symptoms of liver inflammation are observed.^[4]^

Remdesivr contains sulfobutylether beta-cyclodextrin (SBECD) which has no renal toxicities when used in patients with normal renal function (eGFr> 50). However when used in patients with renal impairment renal toxicities were observed. All the existing studies have either used remdesivir in patinets with eGFr > 50 or > 30, the guidelines suggest against its usage when used in patients with eGFr less than 30 and renal parameters have to be regularly monitored.

The kinase inhibitors are proposed as treatments for COVID-19 because they can prevent phosphorylation of key proteins involved in the signal transduction that leads to immune activation and inflammation (e.g., the cellular response to proinflammatory cytokines such as interleukin [IL]-6).^[5]^ Janus kinase (JAK) inhibitors interfere with phosphorylation of signal transducer and activator of transcription (STAT) proteins that are involved in vital cellular functions, including signaling, growth, and survival. Immunosuppression induced by this class of drugs could potentially reduce the inflammation and associated immunopathologies observed in patients with COVID-19.^[6][7]^ Additionally, JAK inhibitors, particularly baricitinib, have theoretical direct antiviral activity through interference with viral endocytosis, potentially preventing entry into and infection of susceptible cells.^[8]^

Adverse effects of baricitinib include infections (typically respiratory and urinary tract infections) and the reactivation of herpes viruses. Additional toxicities include myelosuppression and transaminase elevations. In addition, there may be a slightly higher risk of thrombotic events and gastrointestinal perforation in patients who receive JAK inhibitors. Complete blood count with differential, liver function tests, and kidney function tests should be obtained in all patients before baricitinib is administered and during treatment as clinically indicated. The guidelines have recommend that the use of baricitinib should only be used in conjunction with remdesivir and not as a solo drug. The typical dose recommended is 4mg per day with renal modification in case of renal impairment. ^[16]^

The use of baricitinib and remdesvir combination have shown reduction in the oxygen requirement and help in faster recovery in the trails.^[14]^ Hence it becomes important for us to investigate its impact in our population who are predominantly high risk group and diabetic and its effects on the inflammatory markers.

## REVIEW OF LITERATURE

The ACTT-1 trail a multinational study which studied the use of Remdesivir in the moderate to severe disease have found that there was no difference in the mortality of the patients receiving remdesvir and placebo. They have also found out that there is significant reduction in the oxygen requirement of the patient when on remdesivir and reduced time to recovery seen in moderate disease patients. However they could not find out any significant improvement in the disease condition when they have used in patients undergoing mechanical ventilation and ECMO. They also found out the benefit of remdesivir is there only when it is used within 10 days of symptom onset. ^[9]^

In randomized control trail performed among severe COVID-19 cases in multiple cities of China they have studied the use of remdesivir versus placebo where they have included the patients up to 12 days of symptom onset. They have also found that there is a significant improvement in oxygen requirement and time to recovery was faster when remdesivir was used and no difference was seen when compared with the 28 day mortality. The only limitation of this trail was that the patient also received multiple other immunomodulators additionally like steroids and lopinavir/ritonavir which acted like a confounder in the analysis. ^[10]^

The landmark WHO solidarity trail which was performed in over 11,000 study participants compared the efficacy of the use of remdesivir 10 doses and the placebo have found that there was no difference in the recovery times or the mortality rates in the study populations. They have found out that the incidence of mechanical ventilation in remdesivir group was infact higher than that of the placebo group being 10.8% and 10.5% respectively. ^[11]^

In a randomized control trail which was performed on 592 participants using remdesivir for 5 days in one group and regular standard of care in one group found out that there was a difference in clinical condition of the two groups and the remdesivir group had better discharge rate however the percentages are not wide apart from both groups. ^[12]^ In another study where they have studied regarding the duration of treatment with remdesivir. They have given remdesivir for 5 days in one group and 10 days in other. They have found out that there was no difference in the clinical outcomes when given for 10 days and 5 days. ^[13]^

The ACTT-2 trial which was performed in over 1000 participants had studied the use of remdesivir with baricitinib versus remdesivir with placebo. They had used remdesivir for 10 days and baricitinib 4mg orally for 14 days. They had included the patients who are moderate to severe disease. They have found out that the recovery rate is faster in remdesivir and baricitinib group. The most significant was the reduction in the requirement of oxygen requirement especially those who are on high flow oxygen and NIV with baricitinib combination when compared to remdesivir alone. The recovery ratio was 1: 1.5. However they could not find any significant improvement in the patient requiring mechanical ventilation or ECMO. They have also found out that there was a difference in the 28 day mortality rates, baricitinib and remdesivir group had lesser mortality and however it was not statistically significant. The adverse effect and the risk of secondary infections was also not high in with the infection rate being 6% and 11% in remdesivir plus placebo and combination with baricitinib arms respectively. The only limitation of this study is that it did not evaluate the efficacy of corticosteroids when used with baricitinib . ^[14]^

In our setting **where** we use corticosteroids as the drug of choice in hypoxic patients based on the evidence and mortality benefit showed by the recovery trail^[15]^, **where** there is increase demand for oxygen and medical supplies due to a sudden surge of cases, **where** the ACTT-2 trail clearly demonstrates a positive reduction in the use of oxygen when used remdesivir and baricitinib and on accordance to the trails and guidelines such combinations are being used in treatment, it becomes necessary to investigate regarding the clinical profile and its implications on the biochemical markers in an high risk population

### Research Question?

1. Can the usage of baricitinib with remdesivir lead to improvement from oxygen requirement?
2. Can the clinical recovery be co-related with biochemical recovery?

### Aims and Objectives

1. To find the clinical profile of the patients who were given remdesivir and baricitinib.
2. To find out whether clinical recovery is related to the fall in inflammatory markers.
3. To compare the data thus obtained with already published literature to find out the efficacy of the combination therapy.

## MATERIALS AND METHODS

1. Study setting: Manipal Hospitals, Old Airport Road
2. Study duration: 1^st^ May 2021 to 31^st^ May 2021
3. Study design: A retrospective observational study
4. Study population: Patients admitted COVID-19 infection.

• Inclusion criteria:

(a) All the COVID-19 patients admitted under the departments of Internal Medicine and Geriatrics
(b) All the patients who received the combination of remdesivir (total 5 doses) and baricitinib . The criteria used for baricitinib initiation in our setting was :

1. No response in terms of oxygen requirement even after initial doses of remdesivir, steroids and other standard of care.
2. Systemic sepsis ruled out with procalcitonin being negative.
(c) All the patients who are above 18 years of age..
• Exclusion criteria:

(a) Patients whose data is incomplete
(b) Patients who had received other novel immunomodulators.
5. Study outcomes:

• Primary outcomes: To find out the clinical and biochemical profile of the patient who received remdesivir and baricitinib
• Secondary outcomes:

(a) To find out whether clinical recovery is related to the fall in inflammatory markers
(b) To compare the data thus obtained with already published literature to find out the efficacy of the combination therapy.
6. Sample size: This is retrospective study and would include all the eligible patients who are satisfying the inclusion and exclusion criteria.
7. Data collection methods: The data of the eligible participants shall be taken using a preset study proforma [ANNEXURE-1]
8. Ethical clearance: The study has been approved by the institutional ethics committee and the scientific review committee and the clearance from the committees are enclosed in [ANNEXURE-2]
9. Statistical Analysis: The data thus collected using the study proforma shall be entered in a spreadsheet using Microsoft Excel and will be analyzed using standard statistical techniques.

## RESULTS

The study was started after obtaining the ethics committee and other necessary clearances. The data of all the patients who received remdesivir with baricitinib were scrutinized on the basis of inclusion and exclusion criteria listed in the materials and methods and those who met the study were included. A total of 31 patients were included in the study. Using the study proforma the details of the patient were collected from the patient records and through the electronic patient record system of the hospital. All the data thus collected was entered in Microsoft Excel and then analyzed using standard statistical techniques and the results of the analysis are divided under the following headings

### Patient Demography

1. A total of 31 patients were included in the study of which 24 were males and 7 were females.

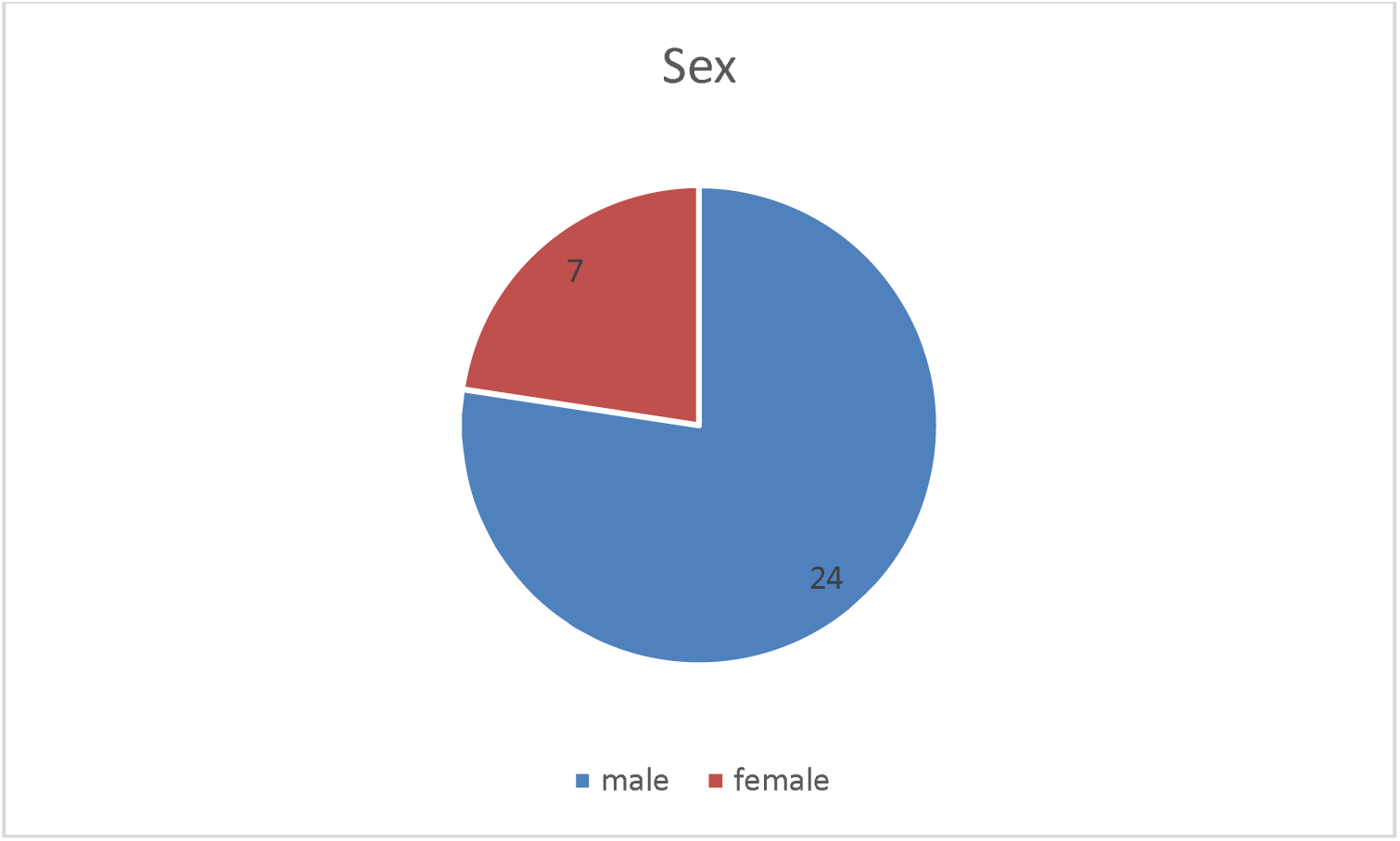
2. Age wise distribution of the patients showed about 13 patient were more than or equal to 59 years. Patients in the age group of 31-50 formed the major part of the study

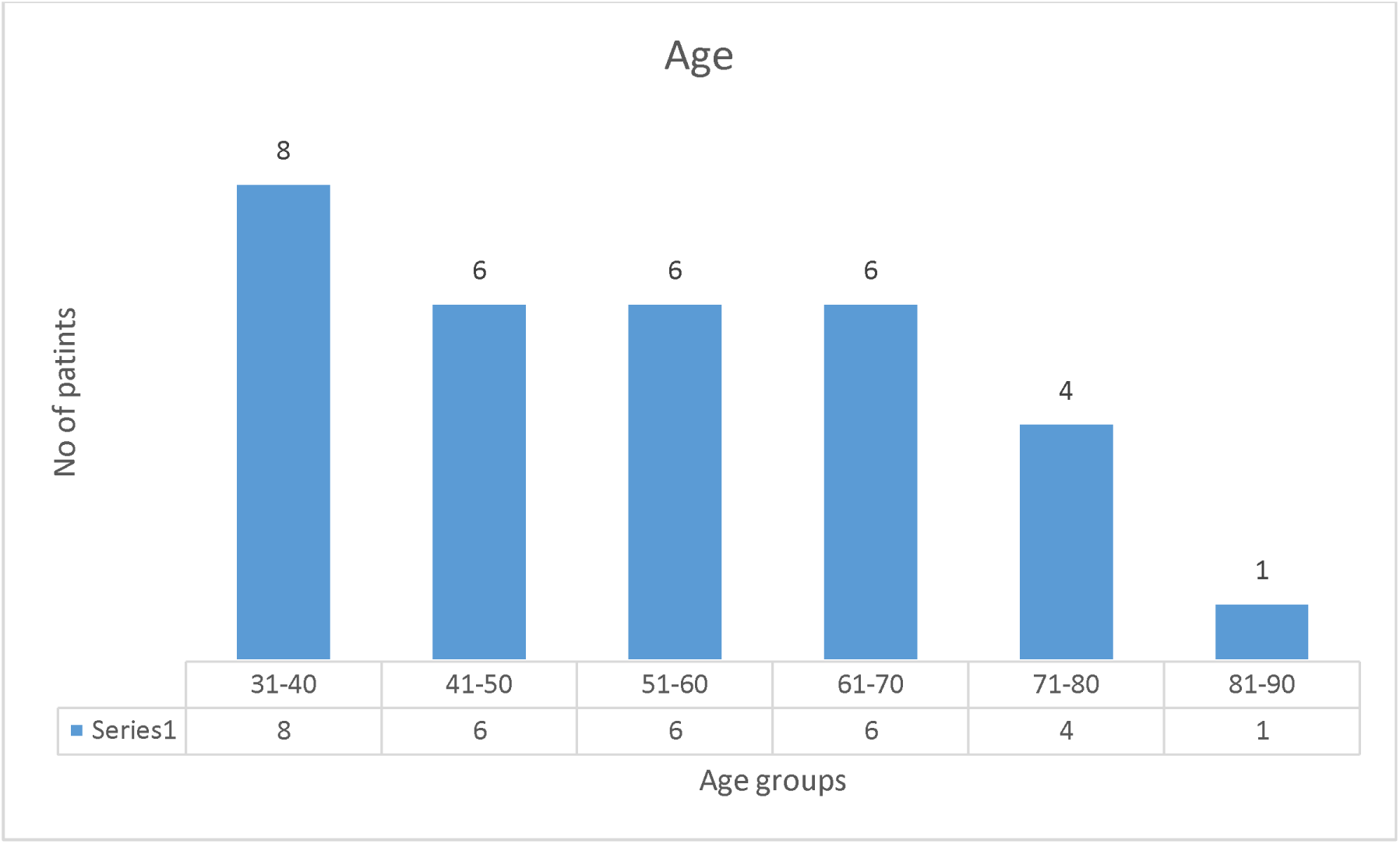
3. It was found that over 50% of the patients were diabetic and nearly 75% of the patients were having co-morbidities

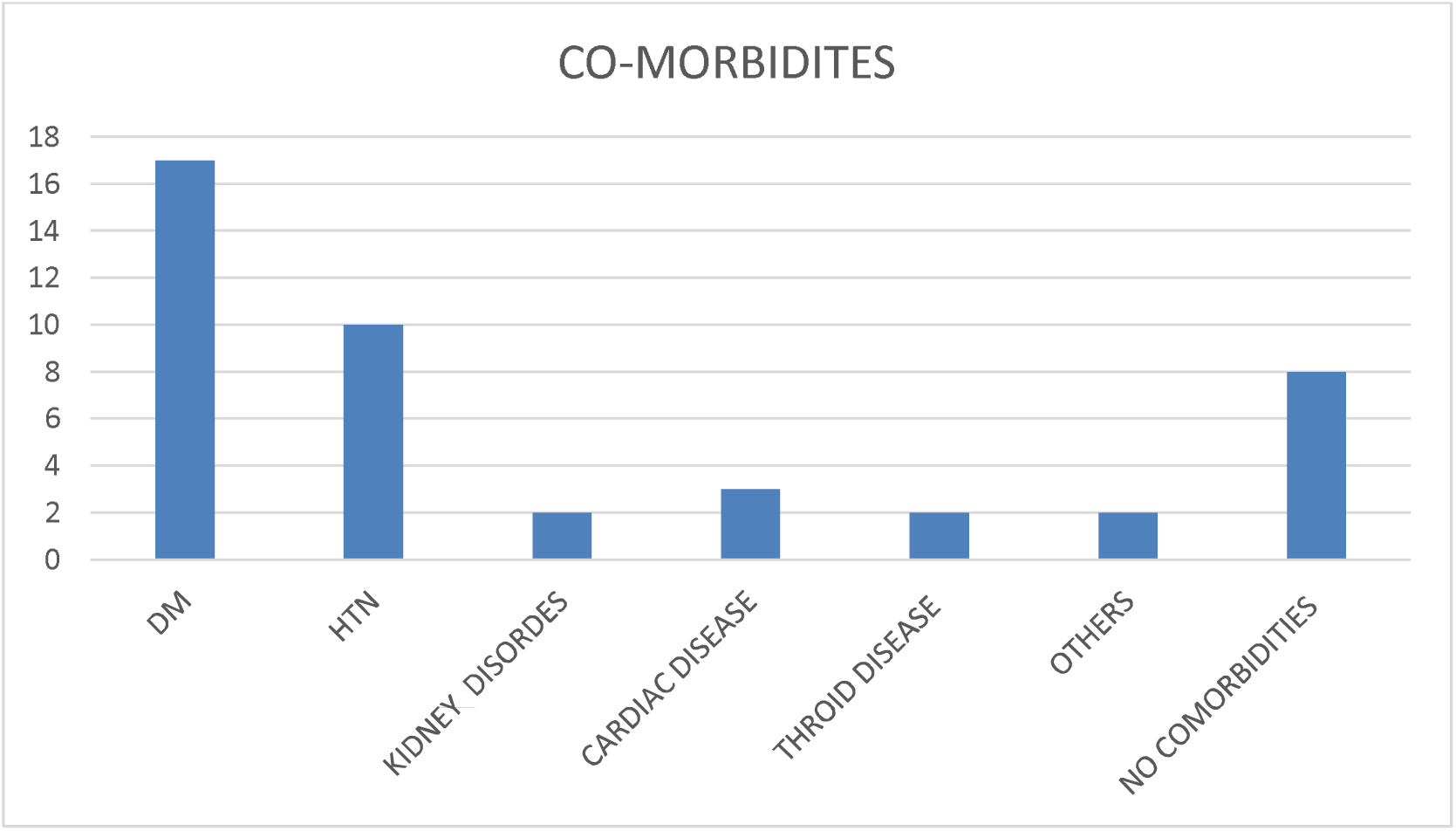
4. All the patients have received remdesivir, baricitinib and steroids along with other supportive measures as per the standard of care of the hospital. Whereas 45% of the patients received methylprednisolone rest received dexamethasone.

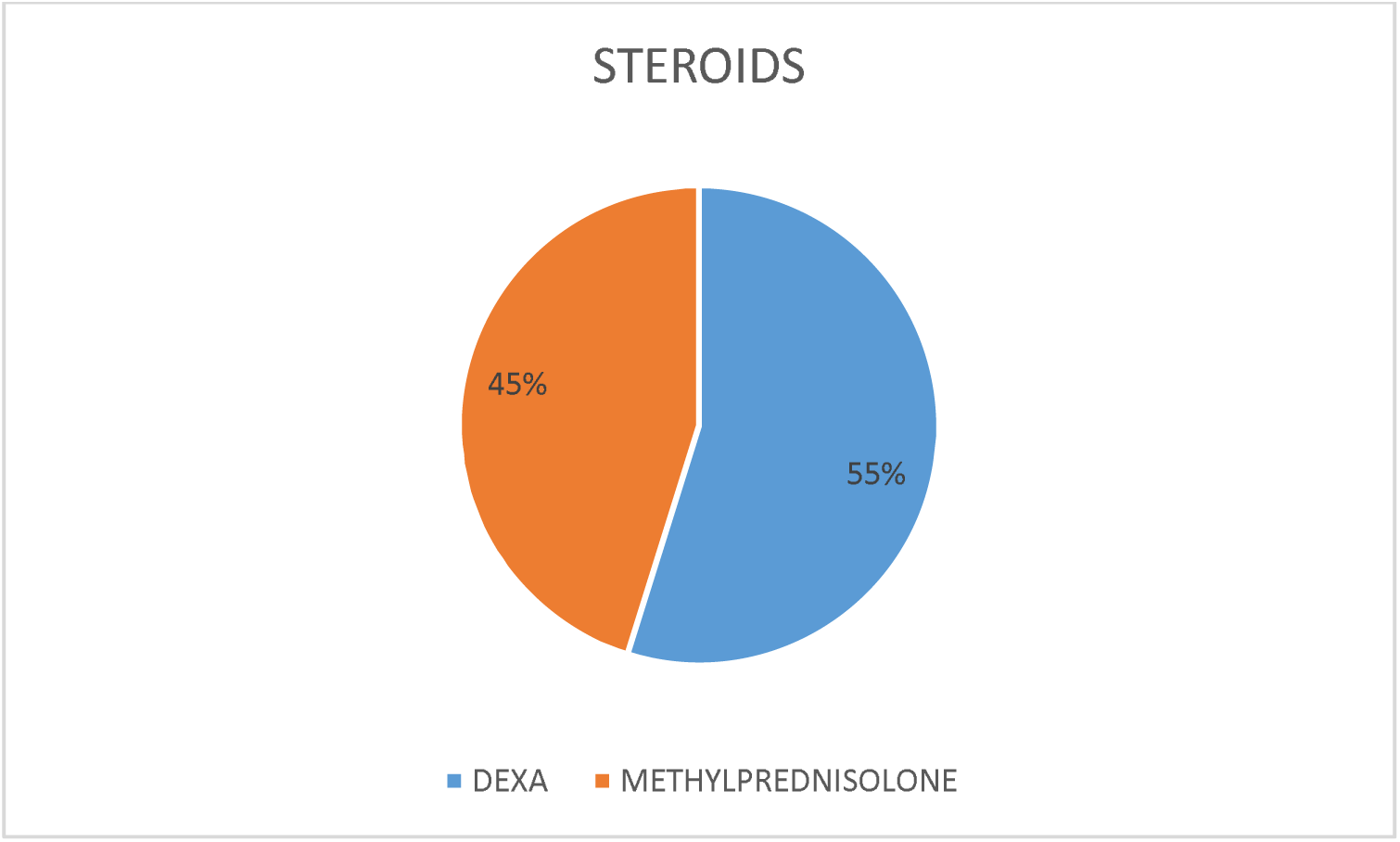

### Oxygen requirement post initiation

1. All the patients were initiated on baricitinib and remdesivir as per the protocols and were frequently monitored. For statistical convenience we have taken the oxygen requirement on day 1, 3, 5 and 7 of the patients. The oxygen requirements of the patients were analysis using ANNOVA and at 95 % confidence levels it was found that there is a significant reduction in the requirement of oxygen p value being <0.05 and r square was 0.997 .
2. Similarly, the oxygen requirement of different sub groups were analysed. When analysed between study population and patients more than 60 years it was found that the oxygen requirement was higher in the older age group and similar reduction in the oxygen requirement was found and it was statistically significant.
3. The comparision between the patients who have comorbidities and those who had no-comorbidities showed that there is similar reduction in oxygen requirement; mean oxygen requirement has come down from 9.2 to 2.3 in subgroup of co-morbidities and almost the patients requirement was down to zero in no comorbidities group ( mean oxygen requirement 5 to 0 ) and the statistically significant.
4. The comparision between survivors and non-survivors showed that there is significant reduction in oxygen requirement with p value less than 0.05 and no significant reduction in the non-survivors and the slope of the curve is almost flat.

Mean Oxygen Requirement

Mean Oxygen Requirement
|  |  | Day 1 | Day 3 | Day 5 | Day 7 | P value |
| --- | --- | --- | --- | --- | --- | --- |
| Age | All age groups | 8.16 | 6.35 | 4.06 | 1.77 | **<u>1.05E-17</u>** |
|  | >60 years | 10.15 | 8.30 | 5.76 | 3.84 | **<u>2.74E-08</u>** |
| Co-morbidities | Present | 9.21 | 7.3 | 4.6 | 2.3 | **<u>1.04E-14</u>** |
|  | Nil | 5.125 | 3.625 | 2.5 | 0.25 | **<u>0.0026</u>** |
| Outcomes | survivors | 7.42 | 5.42 | 3 | 0.6 | **<u>1.77E-17</u>** |
|  | Non survivors | 15 | 15 | 14 | 12.6 | 0.454 |

### CRP post initiation

1. CRP values of all the study participants was taken of day 1, 3, 5, 7 of initiation and analysed using ANNOVA techniques. When analysed taking all age groups into consideration the mean CRP showed reduction from 83.9 to 32.3 and it was found to be statistically significant with p value of 0.0001 at 95% confidence interval.
2. When ANNOVA was performed on patients above the age of 60 years we have a similar trend of reduction ( 91.43 to 71.26 ) and was found to be statistically significant with p value of 0.0043
3. Comparision between patients having co-morbidites and no co-morbidities done showed higher CRP levels in patients having co-morbiditiies and showed a significant reduction in CRP post initiation when compared to patients having no co-morbidities where they showed reduction in CRP from 59.6 to 38.1.
4. Analysis of the CRP values done for survivor and non-survivor groups showed significant reduction in Mean CRP levels from 79.8 to 26.45 with p value of 0.0002 and is statistically significant. However in the non survivors group the CRP values did not show much reduction infact the mean CRP value was increased to day 2 at 130

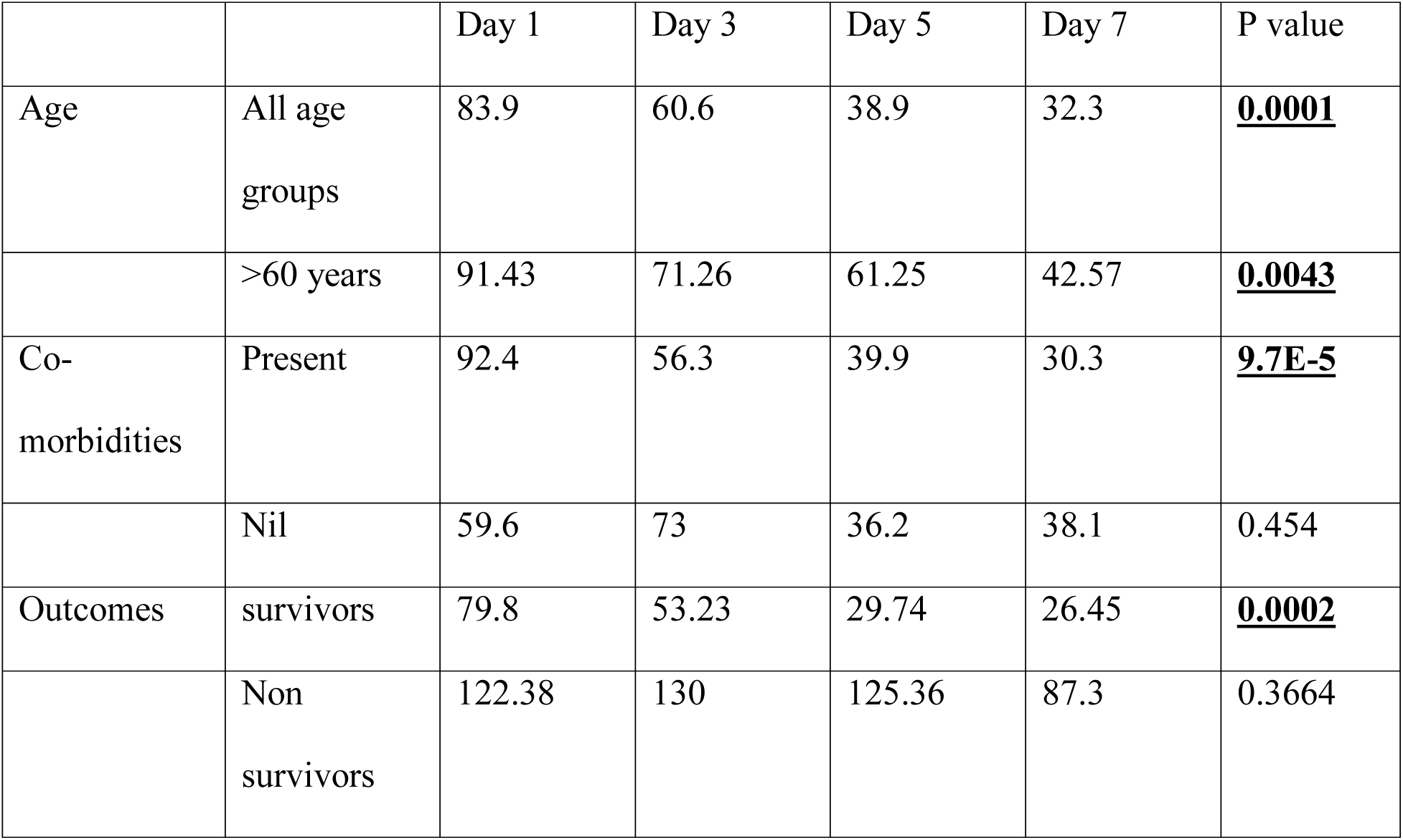
Mean CRP

Mean CRP
|  |  | Day 1 | Day 3 | Day 5 | Day 7 | P value |
| --- | --- | --- | --- | --- | --- | --- |
| Age | All age groups | 83.9 | 60.6 | 38.9 | 32.3 | **<u>0.0001</u>** |
|  | >60 years | 91.43 | 71.26 | 61.25 | 42.57 | **<u>0.0043</u>** |
| Co-morbidities | Present | 92.4 | 56.3 | 39.9 | 30.3 | **<u>9.7E-5</u>** |
|  | Nil | 59.6 | 73 | 36.2 | 38.1 | 0.454 |
| Outcomes | survivors | 79.8 | 53.23 | 29.74 | 26.45 | **<u>0.0002</u>** |
|  | Non survivors | 122.38 | 130 | 125.36 | 87.3 | 0.3664 |

### IL-6

1. The IL-6 values of the patients were taken on days 1, 3, 5, 7 and was analyzed using ANNOVA. The IL-6 values showed a significant reduction from mean IL-6 on day 1 47.5 to mean IL-6 on day 7 being 20.8 and it was found to be statistically significant with p value being 0.004 at 95% and is statistically significant.
2. IL-6 values of age > 60 years also showed reduction from mean values of 51.31 and 32.96 but it was not found to be statistically significant with p value of 0.299
3. Among the patients of having co-morbidities the reduction was seen from a mean value of 483.8 to 21.6 with p value of 0.024 and was statistically significant. While the patients who are not having co-morbidities also showed reduction from 45.3 to 18.6.
4. The analysis done between survivors and non survivors showed significant reduction in mean IL-6 from 48.58 to 29.39 and with a p value of 0.002 was statistically significant. However in the non survivors group the mean showed a rising trend and.
5. **Note: 1 patient with multiple co-morbidities, was initiated on baricitinib and remdesivir however the patient deteriorated and was put on NIV. He went to cytokine storm with IL-6 being >43,000 on day 2 and he passed away. So, for statistical convenience to avoid a single outlier from affecting the analysis, this patient was excluded from the analysis.**

Mean IL-6

|  |  | Day 1 | Day 3 | Day 5 | Day 7 | P value |
| --- | --- | --- | --- | --- | --- | --- |
| Age | All age groups | 47.5 | 31.5 | 27.2 | 20.8 | **<u>0.004</u>** |
|  | >60 years | 51.31 | 56.17 | 45.04 | 32.96 | 0.299 |
| Co-morbidities | Present | 48.38 | 36.75 | 30.5 | 21.6 | **<u>0.024</u>** |
|  | Nil | 45.3 | 17.3 | 18.2 | 18.6 | 0.227 |
| Outcomes | survivors | 49.58 | 29.39 | 27.86 | 21.07 | **<u>0.002</u>** |
|  | Non survivors | 33.43 | 14442.7 | 14414.02 | 14413.14 | 0.454 |

### Neutrophil to Lymphocyte Ratio

1. All the patients NLR was taken on day 1, 3, 5, 7 and was analyzed using ANNOVA. The mean NLR was found to be reducing from day 1 to day 5 and showed increasing trend on day 7 across all sub groups.
2. In all age groups, the mean NLR on day 1 was 13.86 on day 5 was 7.34 while on day 7 it was increased to 12.6. In the age group of > 60 years the mean NLR on day 1 was 17.46 and it came down to day 8 and went up to 14.7 on day 7
3. In the sub groups of co-morbidities, similar trends were found and was not statistically significant.
4. In the sub groups of survivors and non survivors, similar trends were found. Striking rise was found in the non survivors from day 5 to day 7 was 41.24.

Mean NLR

|  |  | Day 1 | Day 3 | Day 5 | Day 7 | P value |
| --- | --- | --- | --- | --- | --- | --- |
| Age | All age groups | 13.86 | 11.11 | 7.34 | 12.6 | 0.139 |
|  | >60 years | 17.46 | 11.98 | 9.06 | 18.04 | 0.306 |
| Co-morbidities | Present | 15.8 | 12.1 | 8 | 14.7 | 0.195 |
|  | Nil | 8.4 | 8.3 | 5.6 | 6.7 | 0.537 |
| Outcomes | survivors | 13.3 | 10.5 | 6.24 | 9.54 | 0.109 |
|  | Non survivors | 19.08 | 16.3 | 17.6 | 41.24 | 0.237 |

### Ferritin

1. All the patients Ferritin was taken on day 1, 3, 5, 7 and was analyzed using ANNOVA. The mean Ferritin showed varying trend on day 7 across all sub groups.
2. In all age groups, the mean Ferritin on day 1 was 911.1 on day 3, 828.9 was while on day 7 it was increased to 1051.14. In the age group of > 60 years the mean Ferritin on day 1 was 720.43 and it came down to day 5, 604.41 and went up to 861.86 on day 7
3. In the sub groups of co-morbidities, similar trends were found and was not statistically significant.
4. In the sub groups of survivors and non survivors, similar trends were found.

Mean Ferritin

**Mean Ferritin**
|  |  | Day 1 | Day 3 | Day 5 | Day 7 | P value |
| --- | --- | --- | --- | --- | --- | --- |
| Age | All age groups | 911.1 | 828.9 | 914.65 | 1051.14 | 0.317 |
|  | >60 years | 720.43 | 829.36 | 604.41 | 861.86 | 0.605 |
| Co-morbidities | Present | 943.1 | 812.6 | 854 | 1062.7 | 0.379 |
|  | Nil | 819.1 | 875.9 | 1089.1 | 1017.9 | 0.211 |
| Outcomes | survivors | 943.5 | 834.48 | 905.19 | 1067.2 | 0.347 |
|  | Non survivors | 608.3 | 777.3 | 1003 | 900.6 | 0.205 |

### D-dimer

1. All the patients D-dimer was taken on day 1, 3, 5, 7 and was analyzed using ANNOVA. The mean D-dimer showed varying trend and did not show any significant decrease in the levels across all the sub groups.
2. However it was found to an increasing trend from day 1 to day3 and slight reduction for day 7, however it was statistically significant.

Mean D-dimer

**Mean D-dimer**
|  |  | Day 1 | Day 3 | Day 5 | Day 7 | P value |
| --- | --- | --- | --- | --- | --- | --- |
| Age | All age groups | 1.73 | 2.55 | 2.5 | 1.97 | 0.353 |
|  | >60 years | 2.61 | 2.48 | 2.95 | 2.56 | 0.679 |
| Co-morbidities | Present | 2.15 | 2.71 | 3.05 | 2.41 | 0.500 |
|  | Nil | 0.55 | 2.10 | 0.93 | 0.72 | 0.546 |
| Outcomes | survivors | 1.64 | 2.47 | 2.24 | 1.78 | 0.451 |
|  | Non survivors | 2.61 | 3.25 | 4.83 | 3.67 | 0.369 |

### Outcome

Out of 31 patients included in the study 27 recovered and 3 patients died of secondary sepsis, and 1 patient had to be shifted to ICU post initiation of baricitinib and is still in the ICU.

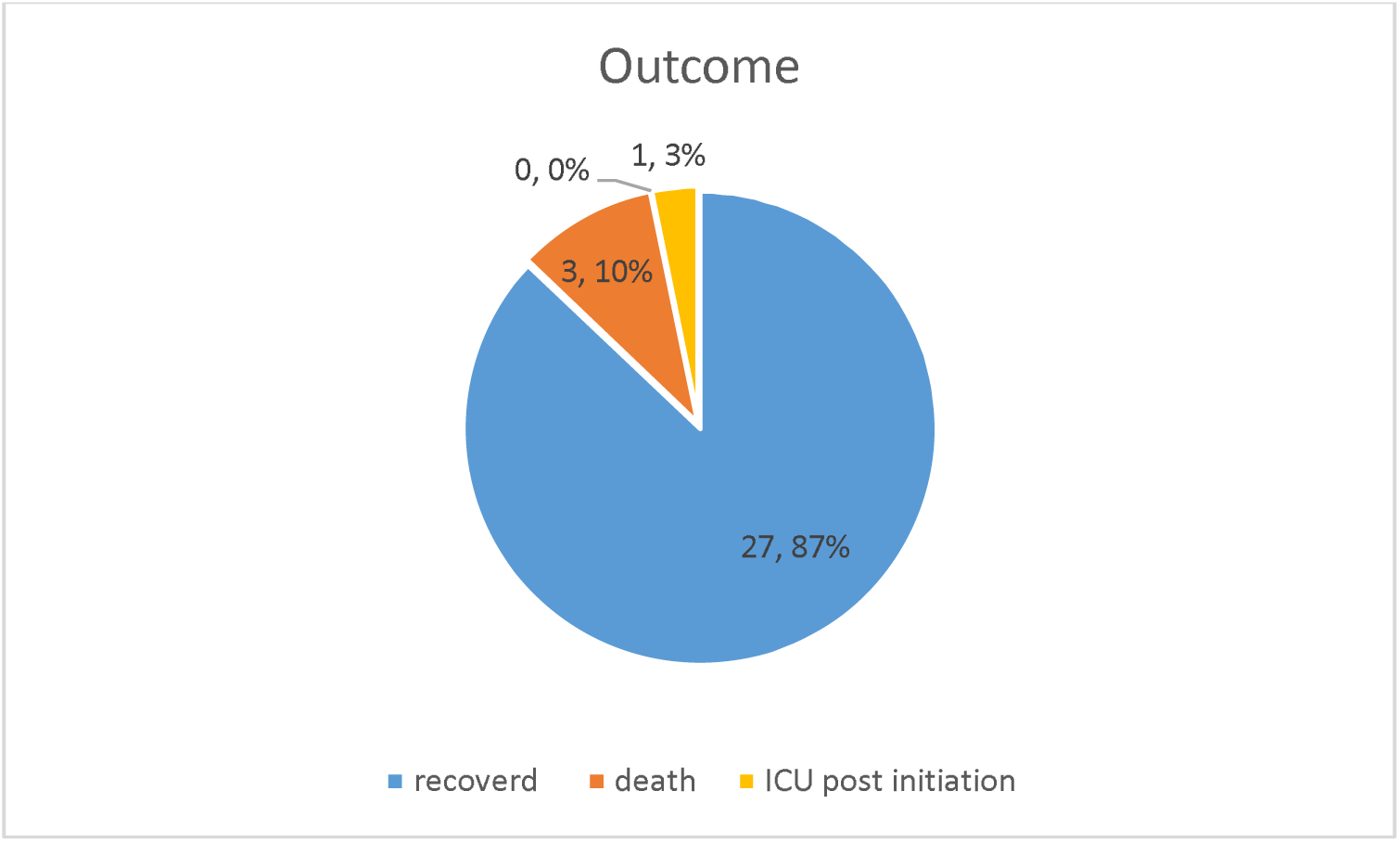

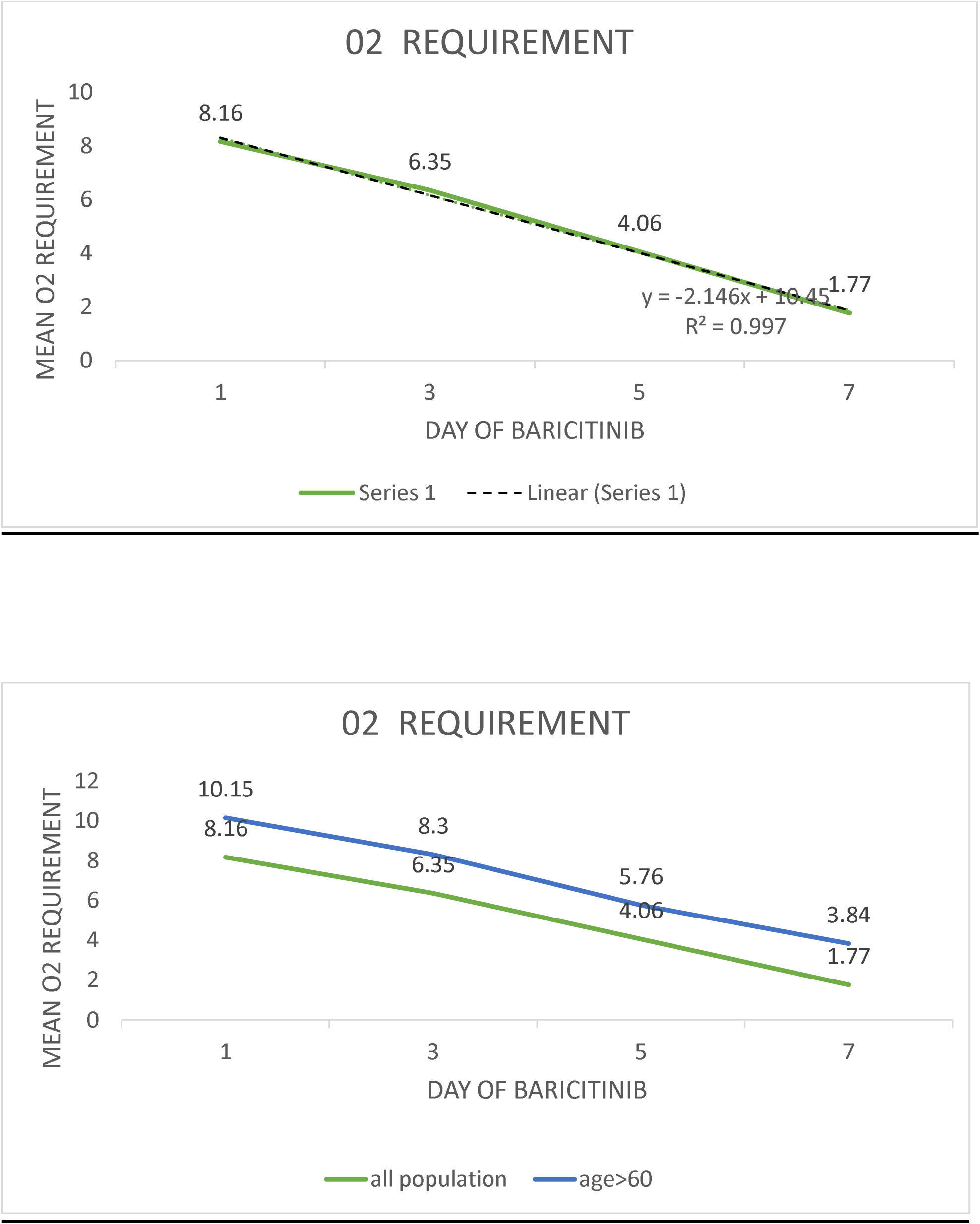

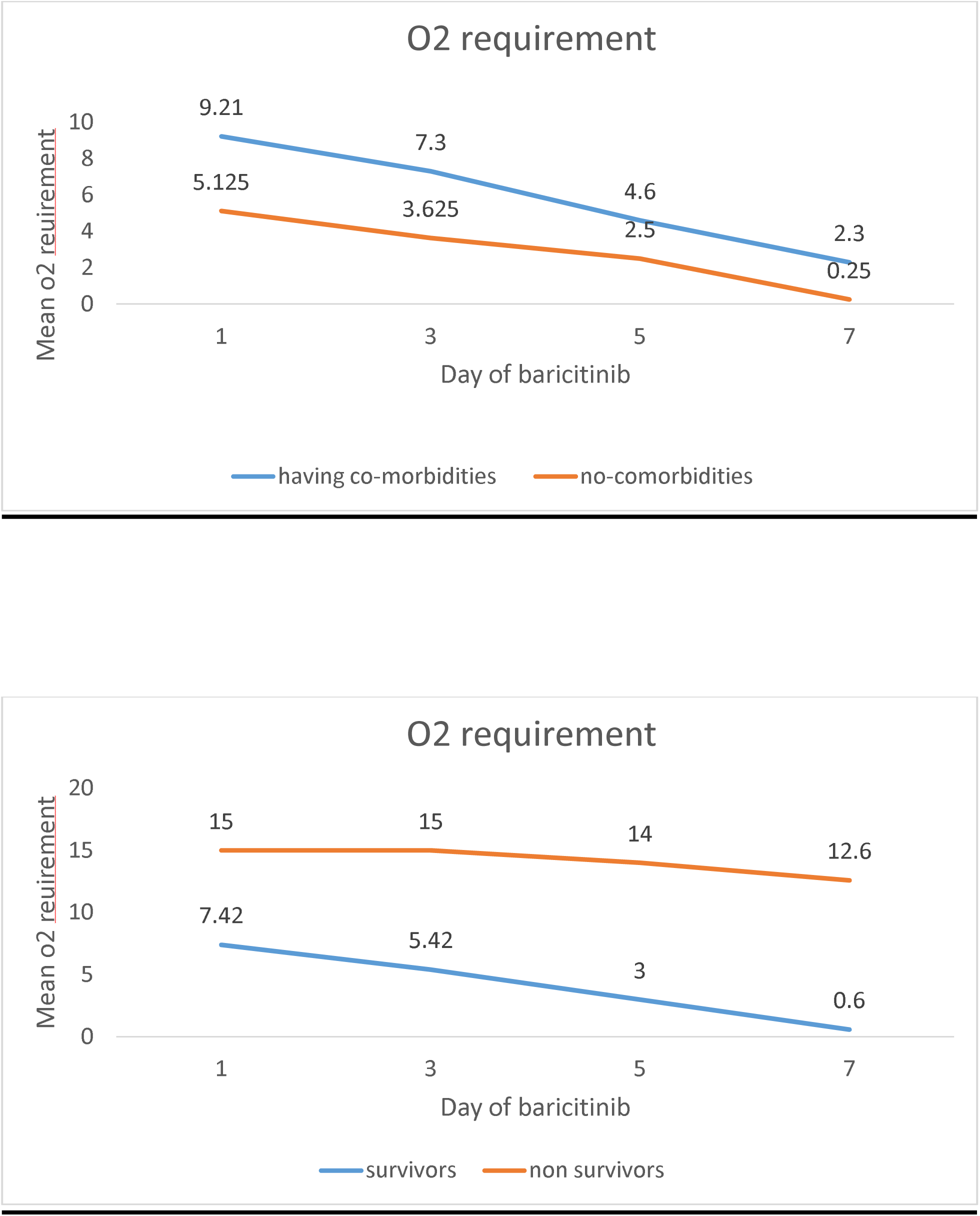

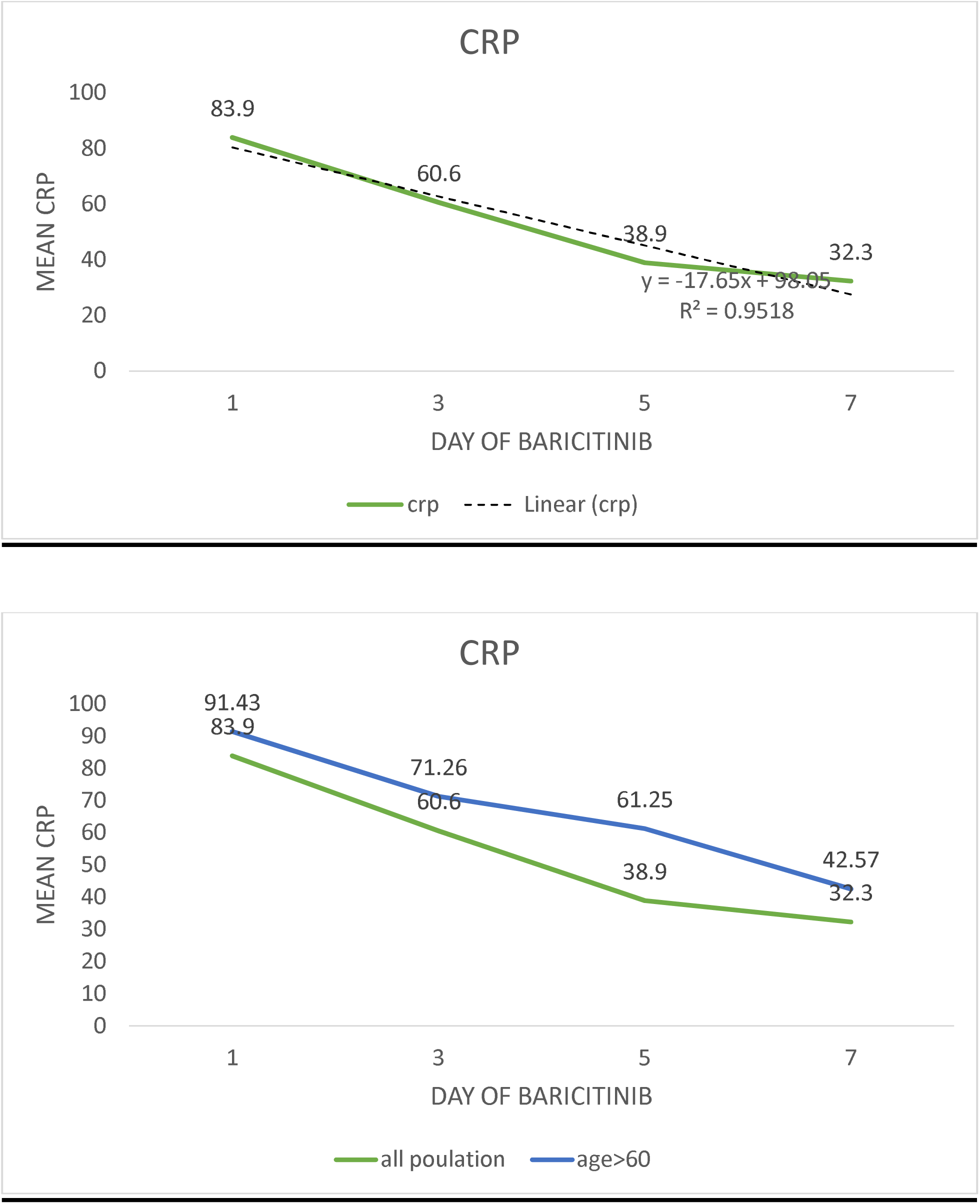

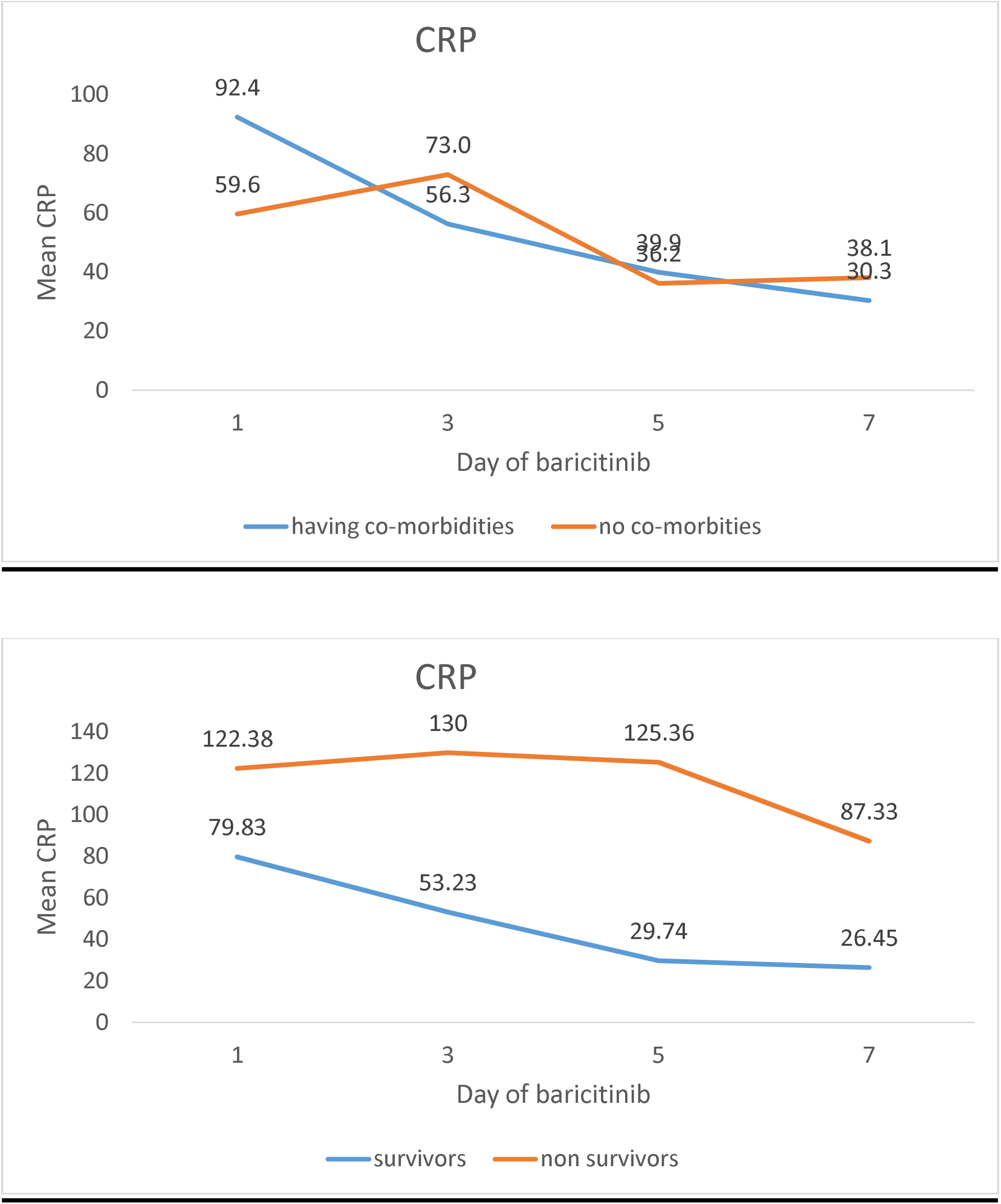

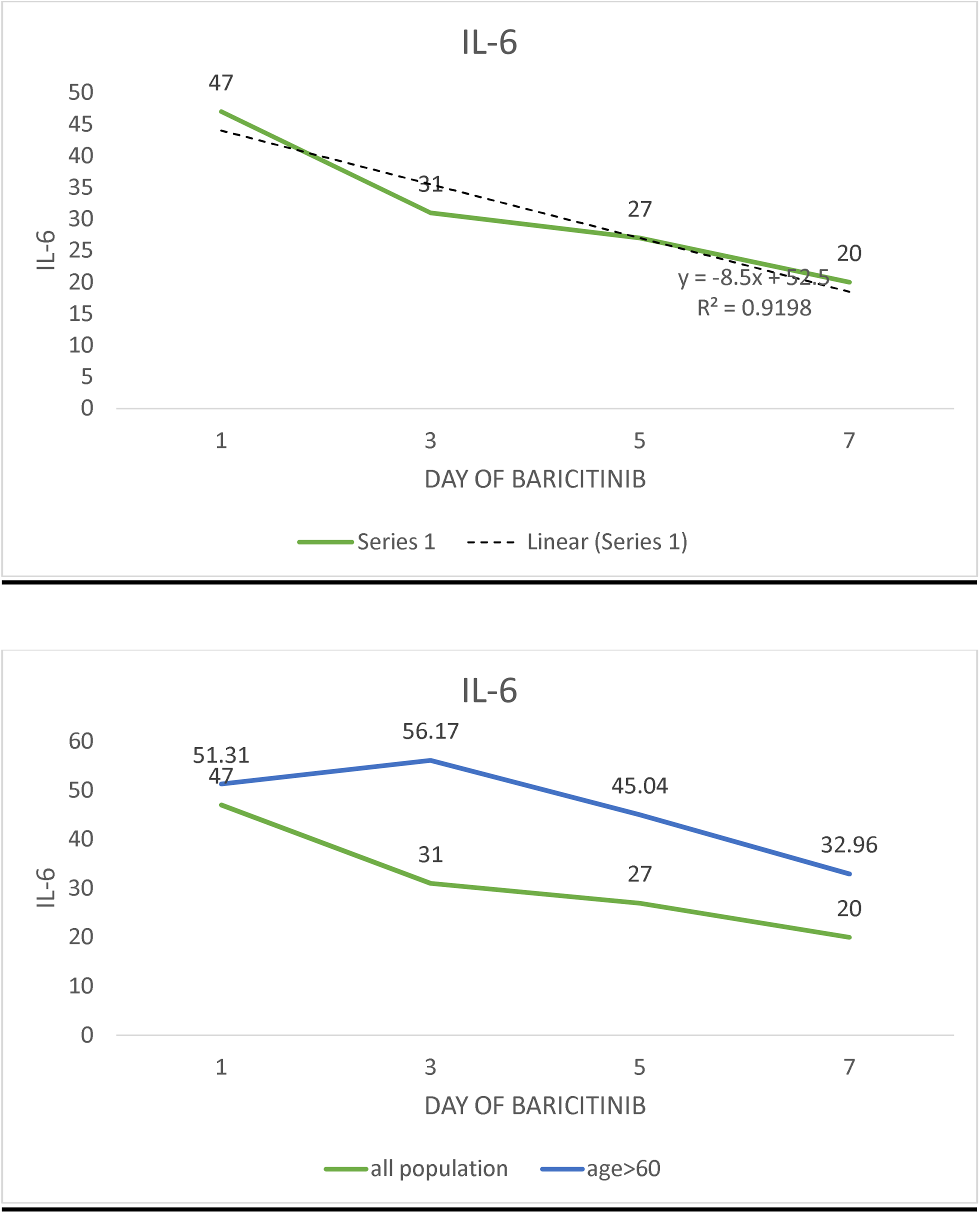

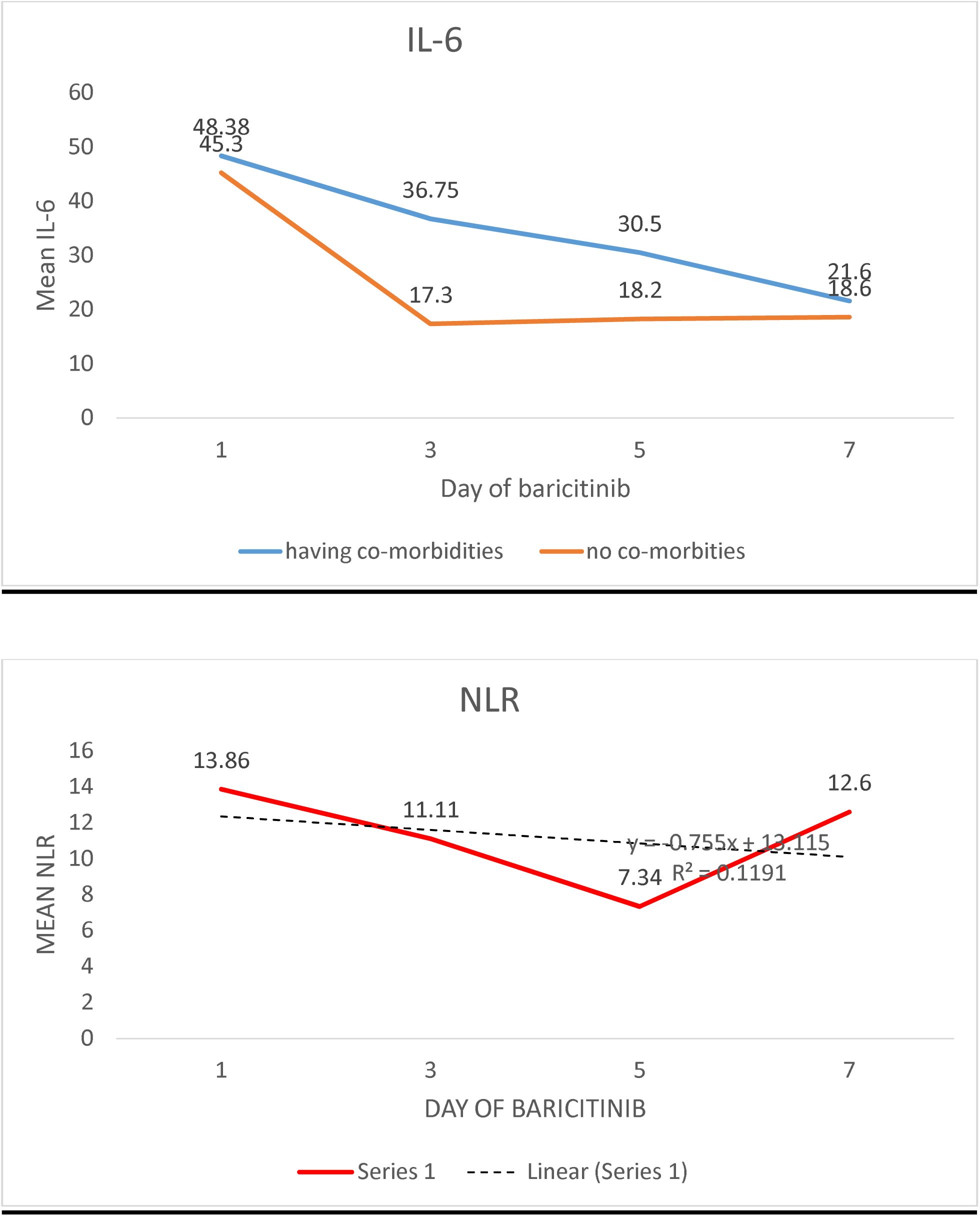

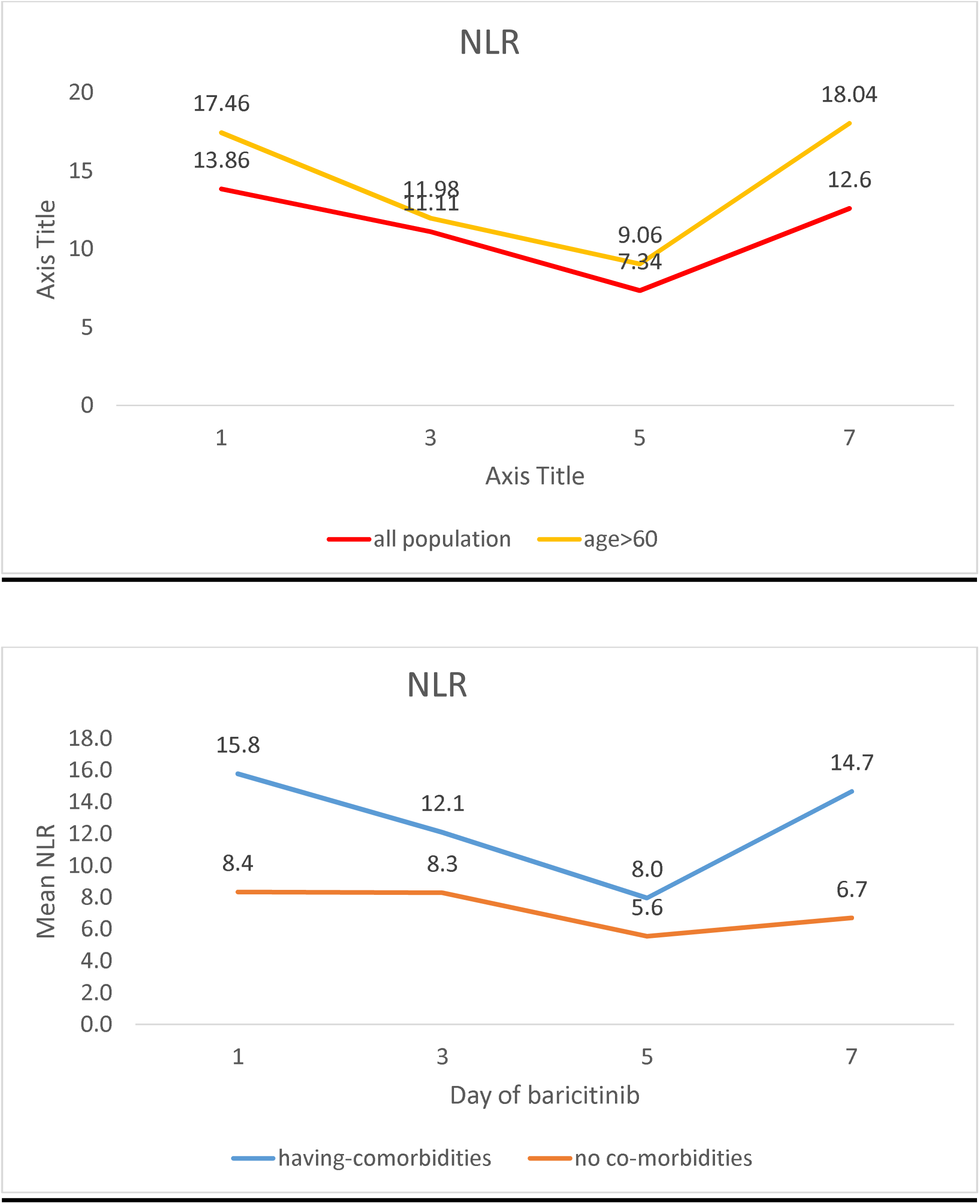

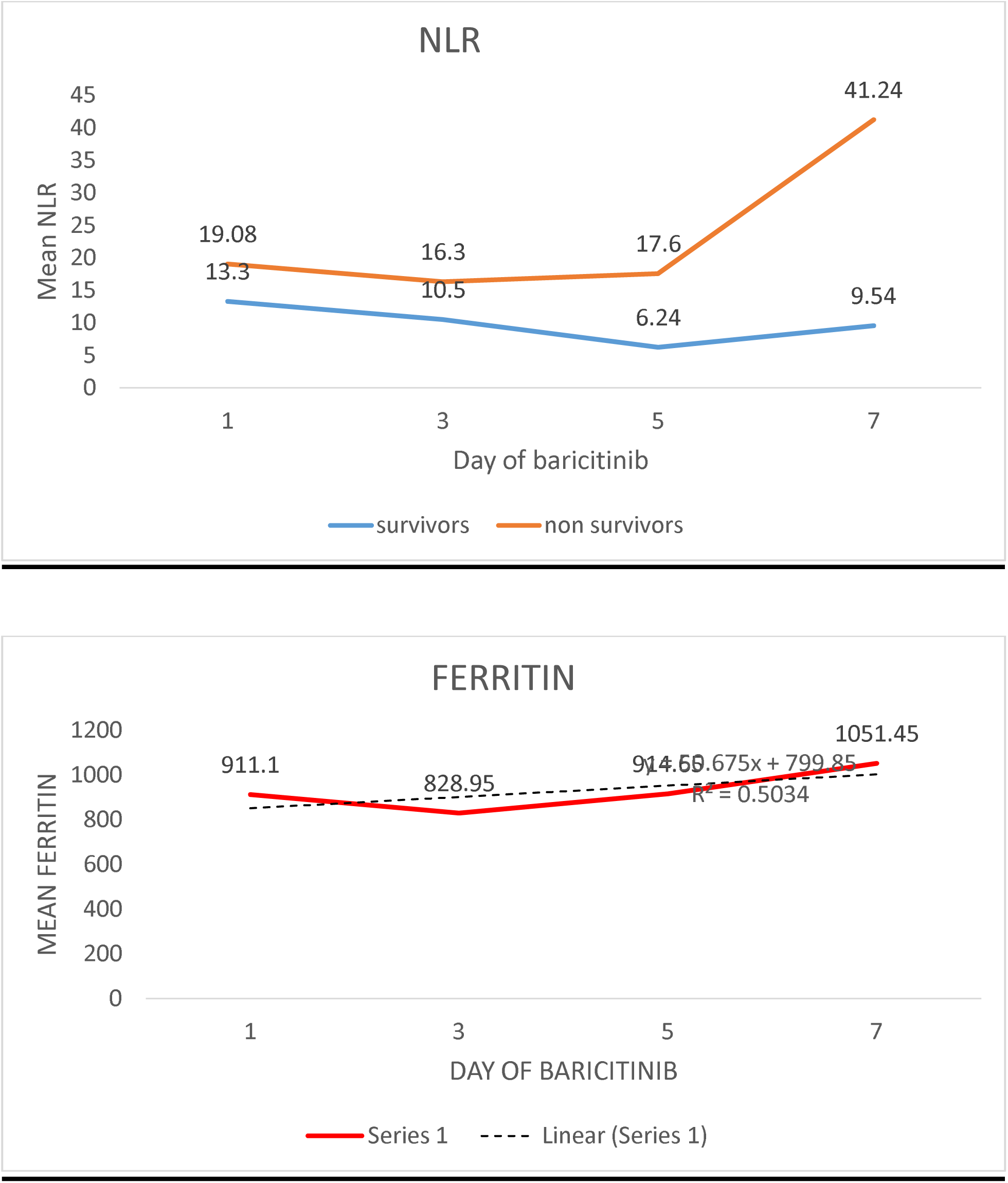

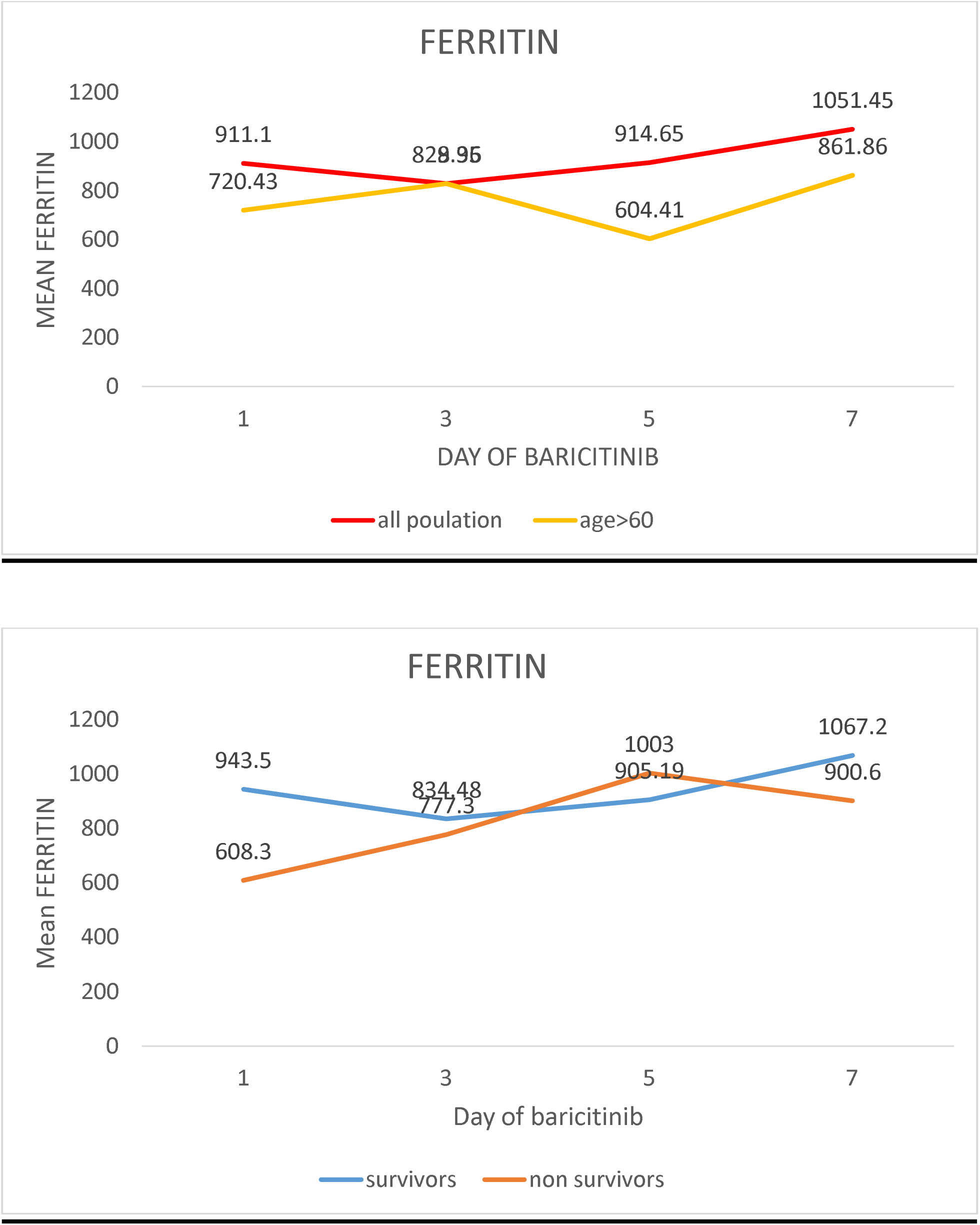

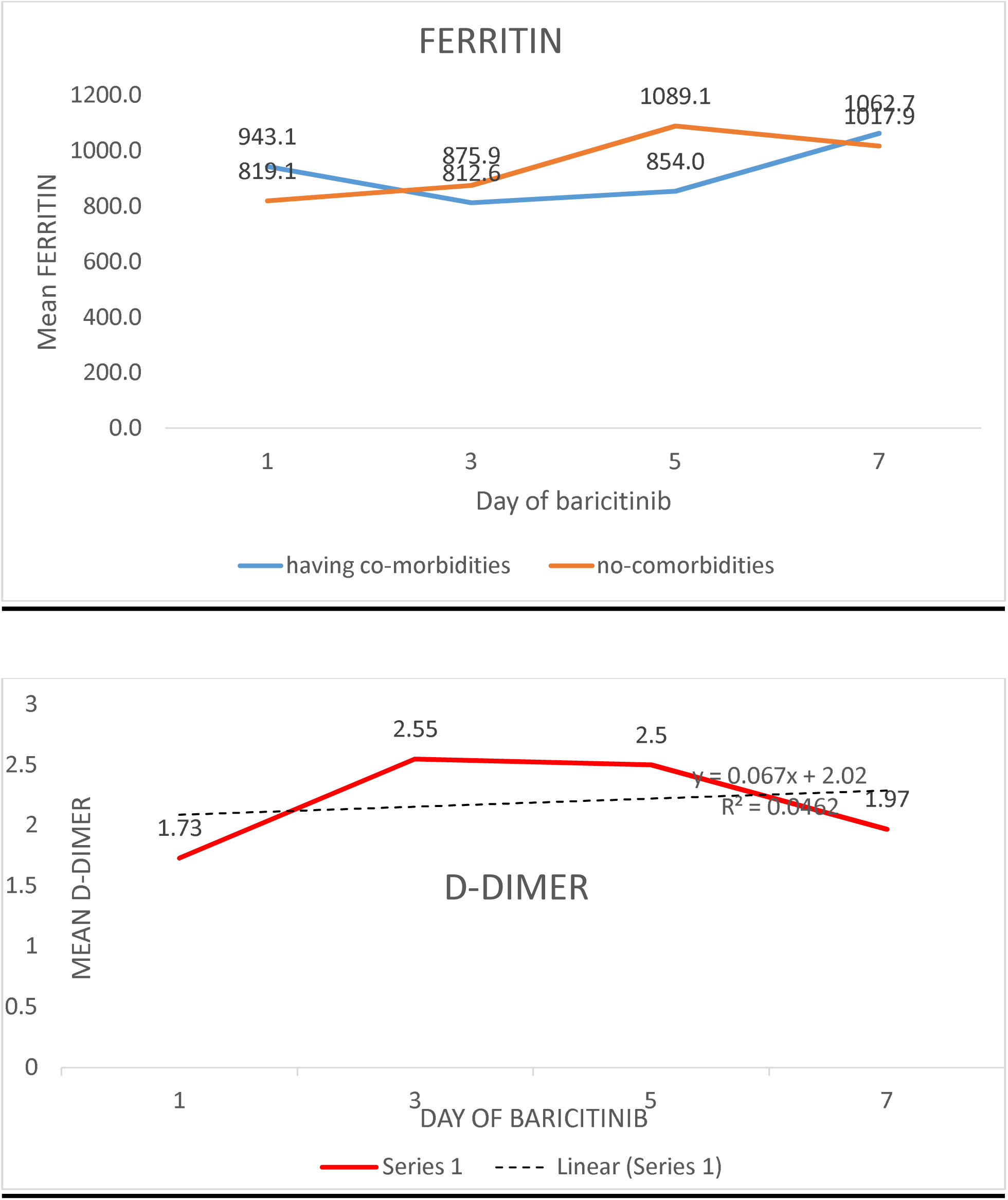

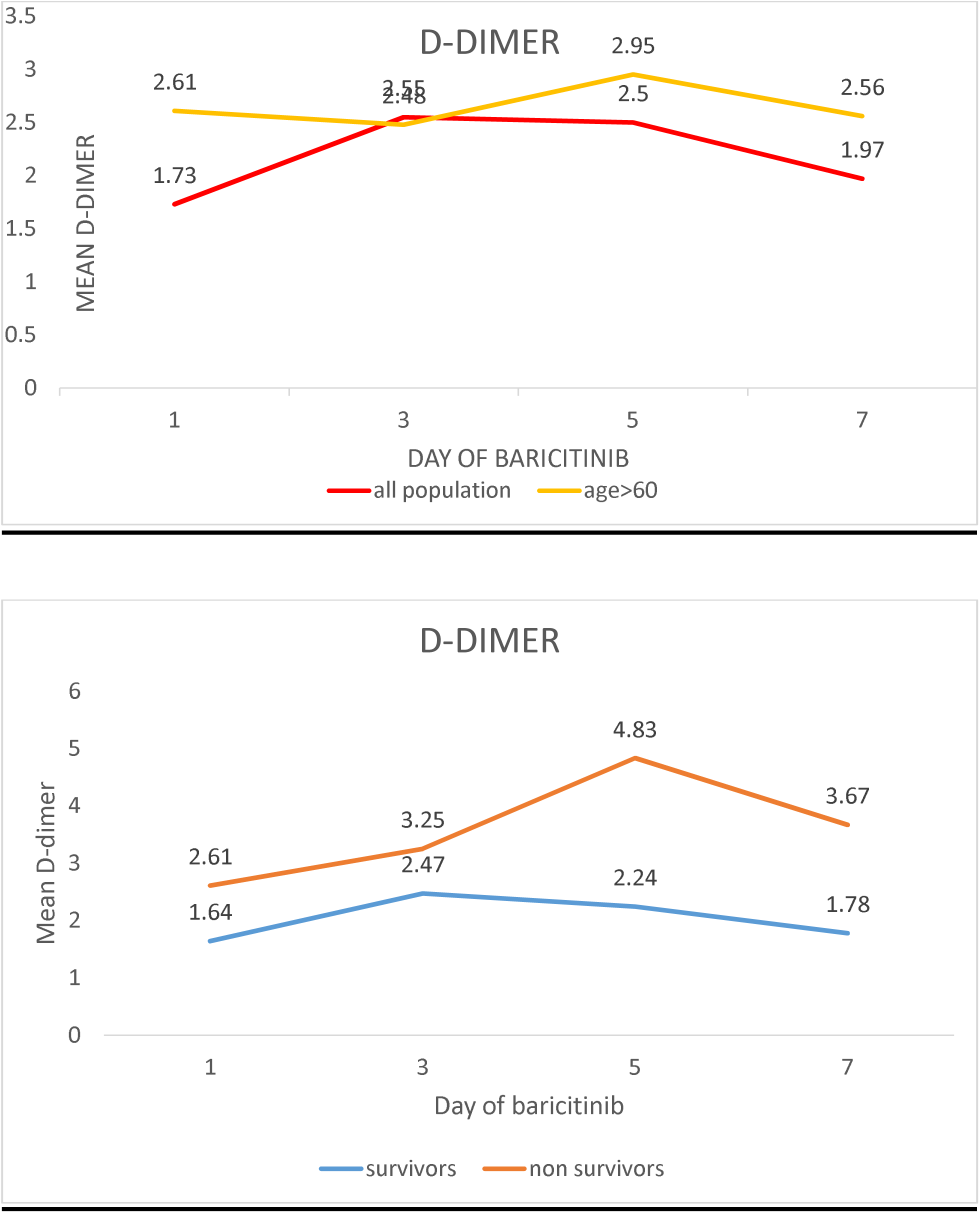

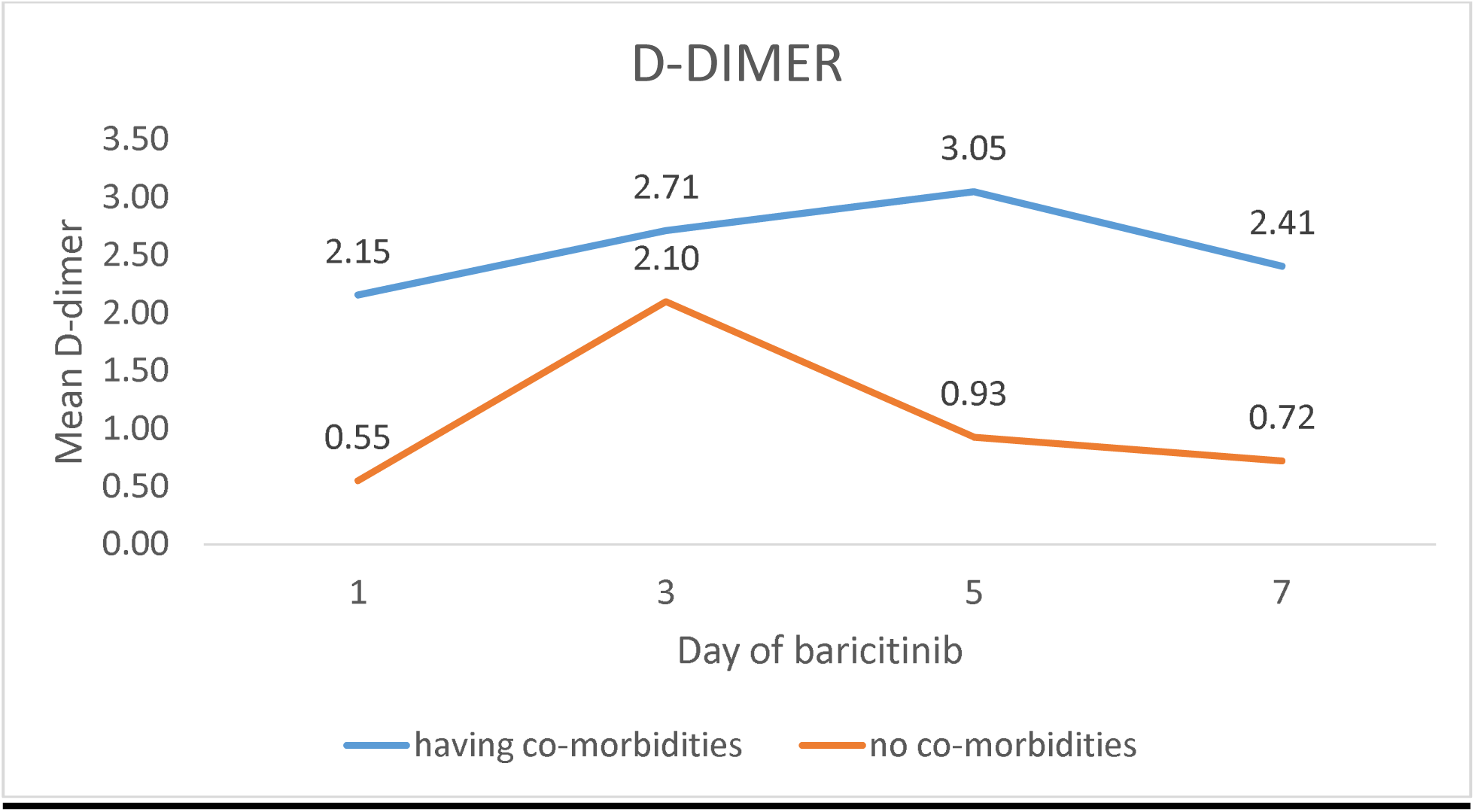

## DISCUSSION

COVID-19 is a single stranded positive sense RNA virus which has spread rapidly in the world leaving a huge impact on the healthcare infrastructure and the economic condition of many condition of the world.^[1]^ Currently, India is experiencing its second wave with the city of Bangalore being most effected. As already stated in the introduction, there was a need to explore newer modalities of therapy which could lead to faster cure of the patients. Among several newer modalities the use of baricitinib and remdesivir was one and was used widely in patients. We have conducted a retrospective observational study to find out the clinical profile and biochemical profile of the patients who received the combination of remdesvrir and baricitinib combination therapy.

The study was conduction in a tertiary care center of south India where there is higher incidence of type 2 diabetes mellitus. This was reflected in our study with the 17/31 of the patients being diabetic. Age wise distribution showed that there was more representation of the people less than 60 years.

The ACTT2 trail which was done to compare the efficacy of remdesivir and baricitinib and remdesivir with placebo showed the mean duration for recovery from oxygen was 7 days.^[14]^ In our study we have similar mean duration of 7 days with a p value <0.05 and it was statistically significant. The increased oxygen requirement was found in the sub group of patients having co-morbidities could be attributed to higher disease burden in them as they are a high risk population and their response to the treatment in terms of reduction in oxygen requirement was much more than those who had no co-morbidities with the r-square values being 0.996 and 0.98 respectively.

The CRP levels of the patients also showed significant reduction with p value of 0.0001. Similar reduction in levels was seen in patients of age> 60. However when we have compared the marker between the sub groups of with and without co-morbidities showed that the subgroup with co-morbidities showed better response to the treatment with a significant p value of <0.05 than the subgroup with no co-morbidities which also showed reduction in the levels but was not statistically significant .

IL-6, which is an important marker for inflammation and predictor of cytokine storm showed significant reduction in the levels, with a p value of 0.004 was significant statistically. When compared the graphs between the sub groups of all population vs age > 60 and the sub groups of patients with and without co-morbidities we concluded that the mean IL-6 levels of the populations age>60 and with co-morbidities were higher than the others similar to the trend in CRP levels.

NLR ratio which is an important prognostic marker in predicting the prognosis and disease severity of the patient ^[18]^ also showed reduction in the levels for the initial days but the ratio was increased towards the end of the treatment. The same trend was observed in all sub groups. This trend of increasing in NLR ratio despite the improving in clinical condition could be attributed to the effect of steroids which the patients were receiving. Shoenfield Y et.al. in their study have stated there is an increase in leukocyte count in patient receiving which increases further as their steroid doses are increased due to various mechanisms.^[19]^

Ferritin which is also an acute phase reactant also increases in COVID-19. It was found that there is no significant change in levels of ferritin in the patient after initiation of baricitinib however we have observed that there was an increasing trend in all the sub groups. Banchini F. et al in their retrospective have found that ferritin is a good acute phase reactant in patients with COVID-19 and its role in predicting the severity and prognosis is not clear. Similarly we have also found that improving clinical condition was not associated with reduction in ferritin.^[20]^

Yao Y., Cao J. et al have established the importance of d-dimer in COVID-19, its importance in predicting the severity and mortality. ^[22]^ In our study we have found that the d-dimer of the patients who are initiated on baricitinib and remdesvir combination had no positive co-relation with the improvement in clinical condition. In the initial few days the d-dimer levels infact had increased and then reduced later. Similar trend was observed in all the sub groups. In the sub group of non survivors the mean d-dimer levels were higher than the survivor group with the highest mean in survivor group being 2.24 and the lowest mean d-dimer in non-survivor group at 2.67

In the ACTT-2 trail, it was concluded that the mortality rate of the group who received the combination of 5.1% and the combination was statistically significant in reducing the mortality among the patients who were receiving high flow oxygen or non-invasive ventilation. The COV-BARRIER study which is in its phase 3 which is being conducted in over 1000 participants with a primary endpoint of prevention of progression of the disease to requirement of high flow oxygen or NIV did not meet its primary outcome as it was not statistically significant, The mortality rate in this study was 8.1% . ^[22]^

The mortality rate in our study is 9.6%. All the three patients who died had co-morbidities and were above 70 years of age. They were all initiated on baricitinib when on high flow nasal oxygen or non-invasive ventilation. While one patient progressed to cytokine storm within 4 days of initiation and died, the others recovered from covid but died due to secondary sepsis. Among the study participants, only three members were on high flow nasal oxygen on non-invasive ventilation at the time of initiation and all of them died due to various reasons while others who were on oxygen support by other means recovered.

The side effect profile of baricitinib combination was found to be 25.1% and 5.5 % in the subgroups who had received and not steroids in the ACTT-2 trail. In our study it is at 9.6% with two people getting severe pneumonia and one person getting localized skin abscess which required incision and drainage. There were no other side effects or adverse effects found other participants over a duration of 28 days follow up post initiation.

## CONCLUSION

The ACTT-2 trail has proved the efficacy of the use of remdesivir and baricitinib combination in COVID-19 patients in reducing the oxygen demand of the patients and also in reducing mortality. In our study we have also found similar results that the combination was helpful in reducing the oxygen demand. However this is a small duration study with a small population and this requires a large scale study for better understanding of the effects.

## LIMTATIONS

It is a single centre study, small duration of 1 month and small study sample of 31 patients. Moreover the effect of corticosteroids have not been accounted for in the study.

## Data Availability

the proforma used for the study and the data sheet shall be made available of required

